# Investigation of neurobiological responses to Theta Burst Stimulation during recovery from mild traumatic brain injury (mTBI)

**DOI:** 10.1101/2022.06.17.22276482

**Authors:** Hannah L. Coyle, Neil W. Bailey, Jennie Ponsford, Kate E. Hoy

## Abstract

The ability of the brain to recover following neurological insult is of considerable interest in mild traumatic brain injury (mTBI) populations. To date, a limited amount of research has examined changes in brain function over time following mTBI. Investigating whether non-invasive brain stimulation (NIBS) can modulate neurophysiology and cognitive performance is particularly relevant for therapeutic targeting post injury. The purpose of the current study was to investigate the neurobiological effects of a single session of intermittent theta burst stimulation (iTBS) applied to the dorsolateral prefrontal cortex (DLPFC) in participants with mTBI during recovery. Changes to neurophysiology were assessed with electroencephalography (EEG) and transcranial magnetic stimulation combined with EEG (TMS-EEG). Digit span working memory accuracy was assessed as a marker of cognitive performance. 30 patients in the subacute phase following mTBI (within one month post-injury) and 26 demographically matched controls were assessed. Participants also completed 3-month (mTBI: N = 21, control: N = 26) and 6-month (mTBI: N = 15, control: N = 24) follow up sessions. Cluster-based analyses demonstrated iTBS did not reliably modulate neurophysiological activity, and no differences were found in cognitive performance in either mTBI or control group participants across any of the assessment time points. The factors that may have contributed to our results are unclear, and possible limitations to our experimental design are discussed. Our findings highlight additional research is required to establish the effects of iTBS on plasticity and cognition in a mTBI population prior to therapeutic application.

## 1. Introduction

The brain’s capacity to recover and adapt following neurological injury has increasingly been recognised (Nudo, 2013). Recovery following mild traumatic brain injury (mTBI) is of particular interest, with mTBI affecting approximately 42 million people worldwide annually (Gardner & Yaffe, 2015). Cognitive, behavioural and affective symptoms following injury have been linked to mTBI pathophysiology (Katz, Cohen, & Alexander, 2015). Although the acute effects of injury are well established (MacFarlane & Glenn, 2015), significant controversy surrounds the prevalence of persistent symptoms (McInnes, Friesen, MacKenzie, Westwood, & Boe, 2017). Most individuals report a return to pre-injury functioning within 90 days (Rohling et al., 2011). However, a subgroup of individuals report persistent symptoms months to years’ post-injury (Bigler, 2008; Sterr, Herron, Hayward, & Montaldi, 2006). Long lasting symptoms have functional implications, being associated with difficulties engaging in social and work activities and reduced quality of life (Polinder et al., 2018; Stalnacke, 2007). There are currently no treatments that are considered effective for acute (Gravel et al., 2013) or ongoing symptoms of mTBI (Prince & Bruhns, 2017). Because of the shearing forces during the mTBI injury, a complex spectrum of axonal abnormalities known as diffuse axonal injury (DAI) are thought to result in dysregulated structural and functional connectivity (Armstrong, Mierzwa, Marion, & Sullivan, 2016; Eierud et al., 2014; Medaglia, 2017). Associations have been reported between dysregulated connectivity, higher levels of self-reported symptoms and poorer cognitive performance, leading mTBI to be conceptualized as a “disorder of brain connectivity” (Hayes, Bigler, & Verfaellie, 2016) and a “disconnection syndrome” (Coyle, Ponsford, & Hoy, 2018). Specifically, damage to regions that are densely anatomically connected and cognitively relevant, such as the frontal lobes (McDonald, Flashman, & Saykin, 2002), contributes to reduced efficiency of functional networks relevant to cognition (Hayes et al., 2016). For example, working memory (a set of cognitive processes that involve actively maintaining and manipulating information) has been explored with measures of neural activity and behavior following mTBI. In response to increased working memory load, mTBI participants have been shown to increase activation in areas outside typical working memory circuity, suggesting recruitment of additional resources as a possible compensatory response post-injury (Chen et al., 2012; Hillary, 2008). Using EEG, differences in the amplitude of event-related potentials (ERPs) (Bernstein, 2002; Ozen, Itier, Preston, & Fernandes, 2013), increased alpha power (Arakaki et al., 2018) and poor coherence (the temporal dependence of neuronal activity across brain regions) between fronto-parietal regions during verbal and visuospatial working memory tasks has also been demonstrated in mTBI (Cudmore, 2000; Kumar, Rao, Chandramouli, & Pillai, 2009; Thornton, 2003). Abnormal coherence (Sponheim et al., 2011) and working memory impairments (Chung et al., 2019) have been linked to DAI, highlighting the relationship between neurophysiology and cognition.

The potential of non-invasive brain stimulation (NIBS) for diagnostic and therapeutic purposes in mTBI has been proposed, but not yet comprehensively explored (Demirtas-Tatlidede, Vahabzadeh-Hagh, Bernabeu, Tormos, & Pascual-Leone, 2012). Transcranial magnetic stimulation (TMS) is a powerful method to measure and modulate cortical activity. A rapid time-varying magnetic field is induced using a handheld coil placed over the scalp and when combined with electroencephalography (EEG), the technique provides a measure of cortical activity (for details see Rogasch & Fitzgerald, 2013). The repetitive application of TMS (rTMS), has been shown to modify neuronal activity locally and distally (Wassermann & Lisanby, 2001). This has significant clinical and therapeutic implications, the induction of long lasting plasticity-related changes in cortical activity, yielding promising application in neurological disorders (Huerta & Volpe, 2009). Theta burst stimulation (TBS), a form of rTMS modelled on the way neurons fire in animal models, involves delivering short bursts of 3 pulses at 50Hz, repeated every 200ms (Huang, Edwards, Rounis, Bhatia, & Rothwell, 2005). TBS can be administered intermittently (iTBS: 2 second trains separated by 10 seconds) or continuously (cTBS: either 20 or 40 s of TBS without any interruption). Markers of cortical activity have demonstrated that iTBS commonly induces a long-term potentiation (LTP) like excitatory plastic effect and cTBS induces a long-term depression (LTD) like inhibitory effect. Furthermore, preliminary evidence suggests applying TBS to cognitively relevant areas, such as the dorsolateral prefrontal cortex (DLPFC) modifies neurophysiology and can modulate cognitive performance (Brunoni & Vanderhasselt, 2014). Improvements in working memory and increased synchronisation of task-related theta (Hoy et al., 2015) and current density changes in resting alpha activity (Grossheinrich et al., 2009) have been reported following iTBS in healthy controls. In summary, iTBS can be applied to investigate post injury plasticity and cognitive performance, providing information on how brain changes may affect cognition following mTBI. Improved understanding of post-injury plasticity may also facilitate the development of directed therapies. To date, no research has investigated functional brain connectivity, plasticity and cognitive performance across recovery in a mTBI population.

The primary aim of the present study was to investigate the effect of iTBS on neurophysiology, and secondary aim to investigate the effect of iTBS on cognitive performance, during recovery following mTBI. We chose a backwards digit span task to explore the involvement of working memory circuitry following mTBI. Neurophysiology and working memory performance were examined before and after administration of iTBS across three time points in both mTBI participants and healthy controls. We hypothesised mTBI participants i) would not show a significant cortical response to iTBS in the sub-acute phase post injury (as measured using TMS-EEG) ii) cortical response to iTBS would increase across recovery, and iii) iTBS would result in increased working memory performance across recovery. Although past literature is mixed, we expected iTBS to significantly modulate neurophysiology and cognitive performance in control participants at each time point.

## 2. Methods and Materials

### Participants

58 participants were recruited (30 mTBI, 28 controls) as part of a longitudinal study investigating clinical symptoms, cognitive performance and cortical activity during recovery from mTBI. The current paper reports the findings from the iTBS plasticity challenge section of the larger study (see Coyle et al 2022a,b). The 30 participants with mTBI were recruited from the emergency department and trauma wards of the Alfred Hospital, Melbourne (less than one month post injury, mean days since injury = 19.70, SD = 16.96, range 10-31, mean age at injury = 35.43 years, *SD* = 10.31). 28 participants with no history of TBI (mild, moderate or severe) were recruited, with two participants excluded due to data collection errors (final sample *N* = 26, mean age = 31.65 years, *SD* = 9.06). The groups did not differ significantly on measures of sex, age and pre-morbid intelligence (all *p* > .05), however controls had a higher level of education *(p* = .035) (Table 1). No participants had a history of seizures, psychiatric or neurological illnesses, unstable medical conditions, were pregnant or taking prescribed medication known to directly or significantly influence EEG findings. mTBI was classified as exhibiting an initial Glasgow Coma Scale (GCS) score of 13-15, loss of consciousness < 30 minutes and post-traumatic amnesia (PTA) < 24 hours (Carroll et al., 2004).

**Table 1.**
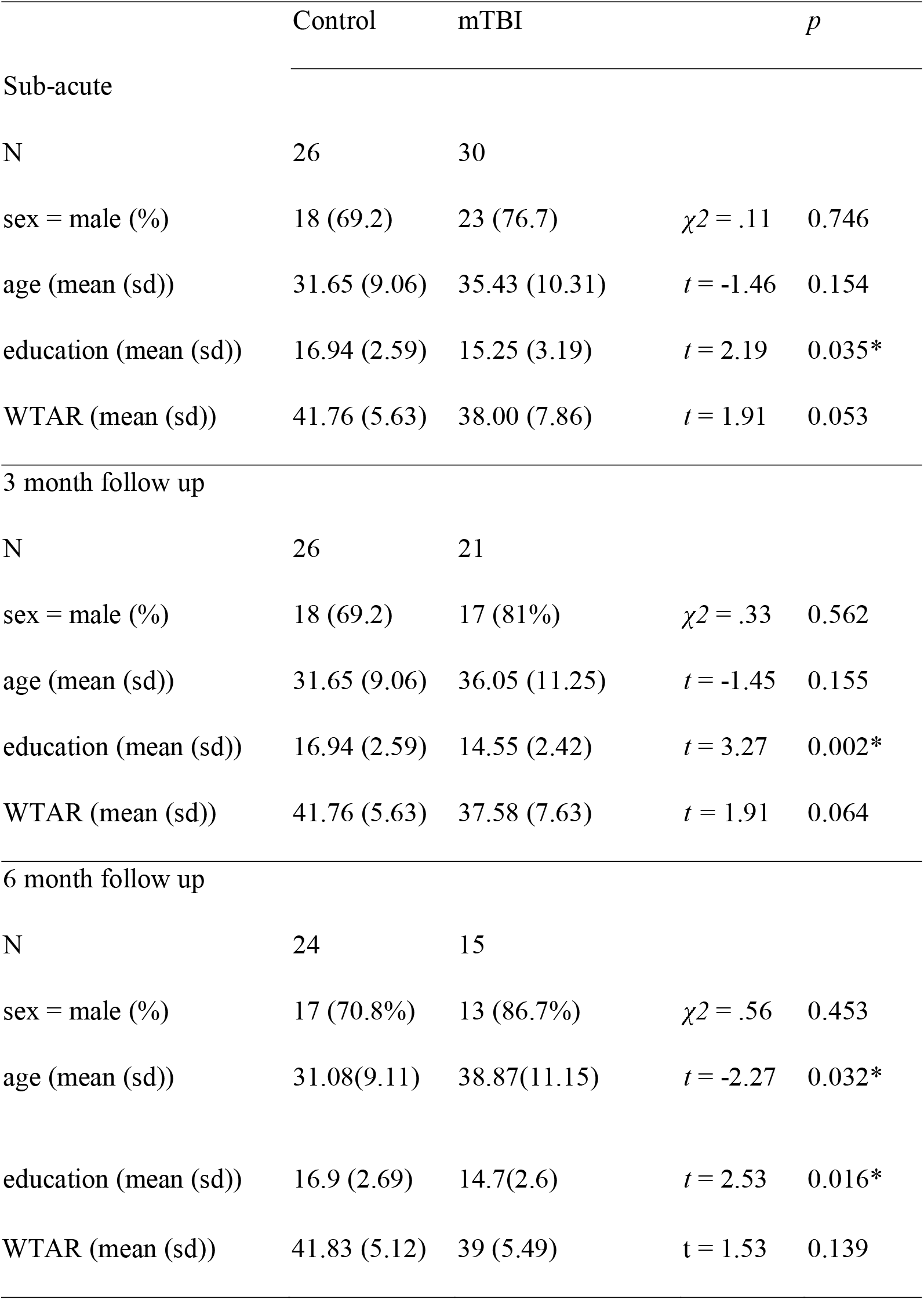
Participant demographics at each time point

All participants provided written informed consent prior to commencement of study procedures. The study received approval from both Alfred Health and Monash University Ethics Committees.

### Procedure

mTBI participants completed assessments within one month post-injury (sub-acute), and at 3-month and 6-month follow up sessions. Attrition resulted in different samples at each time point and demographic and clinical characteristics are reported in Table 1. Control participants completed the same sequence of assessments as a comparison group, as well as to investigate test-retest reliability and cognitive effects in a healthy population. Assessments included recording single pulse TMS-EEG and EEG during working memory before and after administration of iTBS (see Figure 1). Measures are described in detail below.

**Figure 1.**
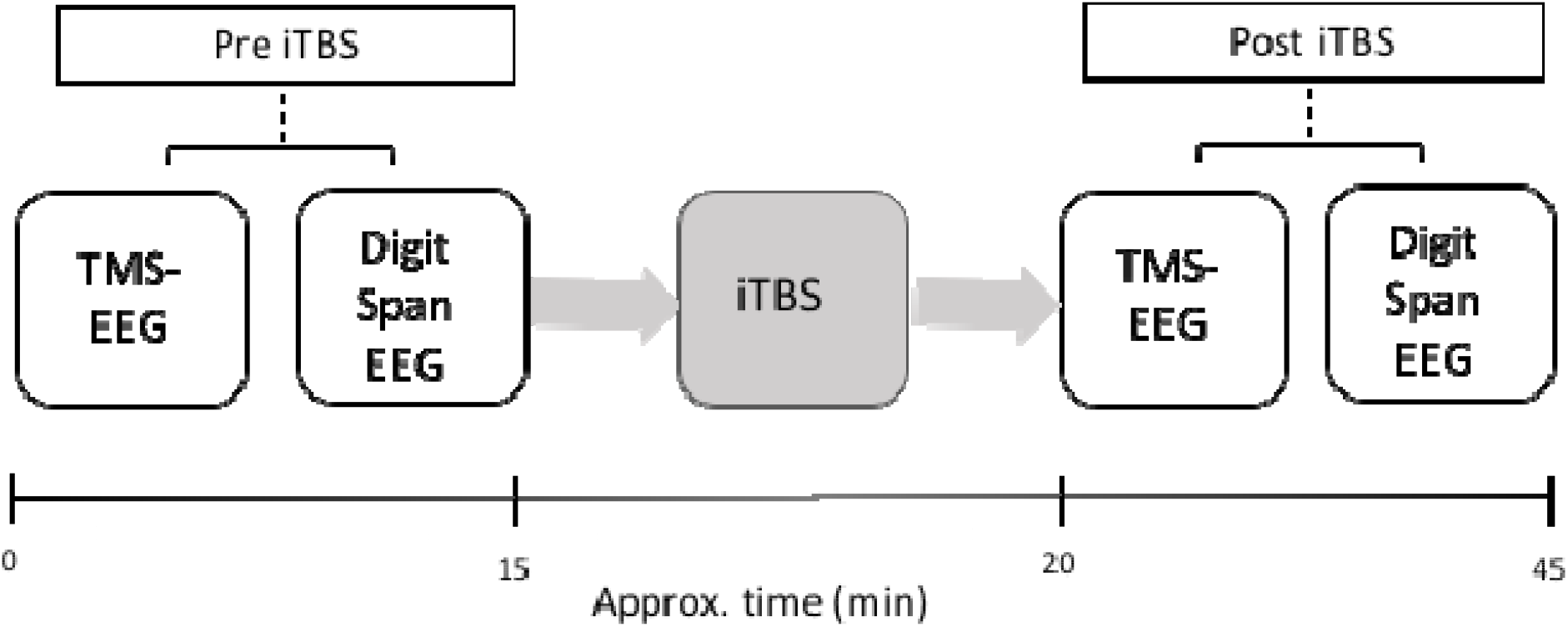
Overview of the experimental procedure

### Neural Measures

#### EEG

EEG was recorded with TMS-compatible Ag/AgCl electrodes and a DC coupled amplifier (SynAmps2, EDIT Compumedics Neuroscan, Texas, USA). Fifty electrodes were used from a 64-channel Easycap EEG cap (AF3, AF4, F7, F5, F3, F1, Fz, F2, F4, F6, F8, FC5, FC3, FC1, FCz, FC2, FC4, FC6, T7, C5, C3, C1, Cz, C2, C4, C6, T8, CP5, CP3, CP1, CP2, CP4, CP6, P7, P5, P3, P1, Pz, P2, P4, P6, P8, PO7, PO3, POz, PO8, PO4, O1, Oz, O2). Electrodes were referenced on-line to CPz and grounded to FPz. All data was recorded with a high acquisition rate (10,000Hz) and low-pass filtered (DC-2,000 Hz) using a large operating window (200 mV). Electrode impedances were kept below 5 kΩ throughout the recording.

EEG was recorded whilst participants completed a computerized version of a backwards digit span task, administered using Presentation software (Version 17.2, Neurobehavioral Systems Inc., Berkeley, CA). Pseudo-randomized sequential auditory digits were presented aurally through earphones in a performance-adapted list length adjustment design. The stimuli consisted of the digits ‘1’ through ‘9’ and digits were not repeated directly in any sequence. The task began with digit sequences consisting of 2 digits and the number of digits in each sequence increased by 1 digit every two trials. The task stopped when the participant had two consecutive errors at any given digit set size. A fixation cross was presented for 1000ms to direct participant’s attention, followed by an interval of 1000ms before the presentation of the first digit. Each digit was presented for 500ms with a 1000ms interval. Following presentation of all digits a recall prompt instructed participants to type their response in using a keyboard and press enter to record their response. To calculate participants raw digit span score one point was given for each successful trial and the total points were summed. Analysis of digit span EEG and task performance at the sub-acute timepoint has been previously reported (Coyle et al., 2022a).

#### TMS-EEG

For the TMS-EEG recordings, single pulse TMS was delivered using a figure-of-eight MagVenture B-65 fluid-cooled coil (MagVenture A/S, Denmark) in a biphasic mode. The EEG cap was applied first, and then the resting motor threshold (RMT) was determined (i.e. by applying TMS over the cap) as the minimum stimulus intensity required to elicit at least three out of five motor evoked potentials (MEPs) > 0.05 mV in amplitude in the relaxed first dorsal interosseous muscles. Single pulse TMS-EEG was administered to the left DLPFC (position F3, international 10–20 system; 45 degrees coil angle; biphasic pulses). During TMS-EEG, participants listened to white noise through intra-auricular earphones (Etymotic Research, ER3-14A, USA) to reduce the influence of auditory processing of the TMS click (Rogasch et al., 2014). The sound level of the white noise was adjusted for each participant until background noise was barely audible. Participants received 100 single pulses at an interval of 4 seconds (with a 10% jitter) at 110% of RMT before and after administration of iTBS.

#### iTBS

iTBS consisted of a burst of 3 pulses given at 50 Hz repeated at a frequency of 5 Hz, with 2 seconds of stimulation on and 8 seconds off, for a total of 600 pulses. iTBS was set at 70% of participant’s RMT. iTBS was delivered to the DLPFC as per single-pulse TMS.

### Data Analyses

All statistical analyses of demographic measures and cognitive performance were performed using R Studio (version 1.1.463) (R Studio Team, 2018). TMS-EEG and EEG data were processed and analysed offline using EEGLAB (Delorme & Makeig, 2004), TESA (Rogasch et al., 2017), FieldTrip (Oostenveld, Fries, Maris, & Schoffelen, 2011), the Randomisation Graphical User Interface (RAGU) (Koenig, Kottlow, Stein, & Melie-García, 2011) and custom scripts on the MATLAB platform (version R2017a).

#### TMS-EEG Analyses

##### TMS-EEG Pre-Processing

TMS-EEG data were epoched around the TMS pulse (−1000ms to 1000ms) and baseline corrected to the pre-TMS pulse period (−500ms to -50ms). Data around the large signal from the TMS pulse (−5 to 15ms) were removed and linearly interpolated. Data were re-referenced to the average and downsampled from 10,000Hz to 1,000Hz. The epoched data from two time points (Pre_iTBS and Post_iTBS) were concatenated and analysed concurrently to avoid bias in component rejection. An initial round of independent component analysis (Hyvarinen & Oja, 2000) (FastICA) was then performed to remove components containing any large residual TMS-evoked EMG artefacts. A bandpass filter (1–100 Hz) was then applied and line noise was removed using a bandstop filter (48–52 Hz). Data was again visually inspected and any remaining noisy epochs were removed. Finally, a second round of FastICA was performed to eliminate any remaining components representing blink, decay and noise-related artefacts. Both rounds of component rejection following FastICA utilised a semi-automated artefact detection algorithm, based on previous research (Rogasch et al., 2014) and using TESA toolbox (Rogasch et al., 2017). Components representing the following artefacts were removed; eye blinks and saccades (mean absolute z score of the two electrodes larger than 2.5), persistent muscle activity (high frequency power that is 60% of the total power), decay artefacts and other noise-related artefacts (one or more electrode has an absolute z score of at least 4).

##### TEP Analysis

Following pre-processing TEP analyses focused on four separate peaks known to occur following stimulation of the prefrontal cortex, the N45, P60, N100 and P200 (Rogasch, Daskalakis, & Fitzgerald, 2015; Rogasch et al., 2014). To investigate group differences and iTBS-induced changes in neurophysiology, cluster based permutation analyses including all electrodes assessed differences in average TEP amplitude (Pre_iTBS vs. Post_iTBS), between groups and within group, for each time point (BL, 3-month, 6-month follow up) separately. Pre-determined time windows for each peak of interest were used for TEP’s [N45 (35-50 ms), P60 (50-70ms), N100 (90-135ms) and P200 (150-240ms)], and data was averaged across time during each window prior to statistical comparisons across the scalp.

First, within-group comparisons were conducted with paired sample t-tests, using raw scores (Pre_iTBS vs Post_iTBS), for each group at each time point separately. T-test designs were used as cluster based permutation analyses implemented in Fieldtrip (the analysis software most commonly used to examine TEPs) are not well suited to 2 × 2 designs (Oostenveld et al., 2011). A primary cluster-forming threshold of *p <* 0.05 and a secondary threshold for family-wise cluster based null hypothesis testing of *p <* 0.025 (two-tailed) was applied with 5000 permutations. Second, between-group comparisons were conducted with independent sample t-tests, using difference scores (Δ), obtained from subtracting Pre_iTBS activity from Post_iTBS activity for each group and each time point.

#### Working Memory EEG

##### Digit Span EEG Pre Processing

Digit Span EEG data were down sampled (1000 Hz), bandpass filtered (fourth-order, zero-phase, Butterworth filter, 0.1-100 Hz) and bandstop filtered (48-52 Hz; to remove 50 Hz line noise). A custom function appended the Presentation .log file to the EEG data to label participant’s responses as ‘Correct’ and ‘Incorrect’. Data were epoched from -500ms prior to digit presentation to 1000ms post stimuli presentation. Digit span EEG files for Pre_iTBS and Post_iTBS were then merged. Automatic artefact rejection was completed, which first checked if more than 3% of epochs included electrodes that varied by more than -250 to 250 microvolts and excluded those electrodes. Next epochs were excluded if they showed a variation of more than 5 SD’s of kurtosis for individual channels, or 3 SD’s for all channels. Lastly, epochs with power within the frequencies 25 to 45Hz that exceeded -100 or 30 dB were excluded (power in these frequencies usually reflects muscle activity). Manual artefact rejection was then completed to ensure the automatic process did not miss significant artifacts, the data being visually inspected to remove epochs with excessive noise (i.e. muscle artefact), and bad channels (i.e. disconnected). AMICA (Adaptive Mixture of Independent Component Analysis) (Palmer, Kreutz-Delgado, & Makeig, 2011) was used to manually select and remove eye movements and remaining muscle activity artefacts. Rejected electrodes were re-constructed using spherical interpolation (Perrin, Pernier, Bertrand, & Echallier, 1989). To ensure data was in the correct form for RAGU analysis, epochs from the Pre_iTBS and Post_iTBS data were averaged separately, data was re-referenced to the average reference and correct responses were baseline corrected to the period from -500 ms to stimulus onset.

Following pre-processing participants were included only if they had provided =>15 artefact free correct epochs for both Pre_iTBS and Post_iTBS time points. Only individuals with sufficient epochs for EEG analysis were included in cognitive analyses (i.e. digit span accuracy).

For statistical analysis of ERP’s, measures of neural response strength (Global Field Power: GFP) and topographical distribution of neural activity (Topographical analysis of variance: TANOVA) were calculated using RAGU. RAGU is a multivariate approach that uses powerful, assumption free, randomisation statistics to analyse multi-channel event related potential (ERP) data (Koenig et al., 2011). RAGU allows for comparisons of overall neural response strength (with the global field power -GFP test). Global brain activity can be described by the global field power (GFP), which is mathematically defined as the root of the mean of the squared potential differences at all *K* electrodes (i.e. *V*_*i*_*(t)*) from the mean of instantaneous potentials across electrodes (i.e. *V*_*mean*_*(t)*) (Lehmann & Skrandies, 1980). A measure of the strength of the electric field over the brain at each point in time, GFP is representative of the global brain response to an event. Local maxima of GFP curve represents instances of strongest field strength and highest topographic signal to noise ratio (Khanna, Pascual-Leone, Michel, & Farzan, 2015). RAGU also allows for comparisons of the distribution of neural activity between group and condition, using topographic analysis of variance (TANOVA).

The TANOVA is a non-parametric randomisation test based on global dissimilarities between electric fields. In contrast to electrode-wise comparisons, the TANOVA computes global dissimilarity of the whole electrical field topographies between conditions or groups and tests for the significance of these topographic differences at each time point (Ruggeri, Meziane, Koenig, & Brandner, 2019). The TANOVA was implemented on the amplitude-normalized maps (GFP = 1), such that the results obtained are independent of variations in the global field strength. The rationale behind this approach is that it enables significant differences between conditions to be attributed to partially different sources of the evoked potential, and not to different strengths of similar source distributions. To protect the results from false positives caused by multiple testing, additional testing checked whether the duration of continuous periods of significance observed in our data exceeded the duration of significant periods in > 95% of the randomised data. This ensured the duration of a significant time period exceeded chance. Five thousand permutations were conducted with an alpha of *p* < 0.05.

##### Digit Span EEG Analyses

Due to the performance-adapted list length adjustment design nature of the task, the number of epochs per participant varied. Only participants with =>15 correct epochs per time point (Pre_iTBS and Post_iTBS) were included in the analysis. This number of epochs was chosen to maximise data inclusions. 7 participants were excluded due to too few correct epochs at the sub-acute timepoint (Control, *N*= 2, mTBI, *N*= 5), 3 participants at 3 month follow up (Control, *N*= 1, mTBI, *N*= 2) and 1 participant at 6 month follow up (mTBI, *N*= 1). The number of available epochs was not significantly different between Pre_iTBS and Post_iTBS conditions for control or mTBI participants at any time point (*p >* 0.05). However, control participants had a greater number of total available epochs than mTBI participants at all time points (*p <* 0.05). This was because mTBI participants had lower digit span accuracy scores, which resulted in fewer available epochs. However, the primary aim was to investigate the within-group effect of iTBS. Prior to statistical analysis, automatic outlier detection using multidimensional scaling in RAGU was used to detect and exclude extreme values. At the sub-acute timepoint this resulted in the removal of 2 participants (Control, *N*= 1, mTBI, *N*=1) and at 3 month follow up 2 participants (Control, *N*= 1, mTBI, *N*=1). The automatic outlier detection is based on an algorithm that uses the Mahalanobis distance among the displayed points to identify cases that are unlikely to be part of the normal distribution (Habermann, Weusmann, Stein, & Koenig, 2018).

### Statistical analysis

To investigate group differences and iTBS-induced changes in the EEG data, a 2 × 2 repeated measures ANOVA design was performed in RAGU with CONDITION (Pre_iTBS vs Post_iTBS) as a within subject factor and GROUP (control vs mTBI) as a between subject factor. To investigate group differences and iTBS-induced changes in cognitive performance (as measured by digit span accuracy), a 2 × 2 repeated measures ANCOVA was performed with CONDITION (Pre_iTBS vs Post_iTBS) as a within subject factor, GROUP (control vs mTBI) as a between subject factor and estimated pre-morbid IQ (WTAR) as a co-variate (to control for the potential influence of pre-morbid IQ). As the study’s aim was to investigate the effect of iTBS on working memory, only interaction effects are reported. Participant attrition prevented the inclusion of time point as an additional within subject factor for the repeated measures analyses (see Table 1 for sample sizes for each group and time point). Instead analyses were conducted separately for each time point (BL, 3-month and 6-month follow up).

## 3. Results

An overview of the experimental protocol is provided in Figure 1.

### TMS-evoked potentials (TEPs)

TEP grand average waveforms for mTBI and control group participants at each time point are shown in Figure 2.

**Figure 2.**
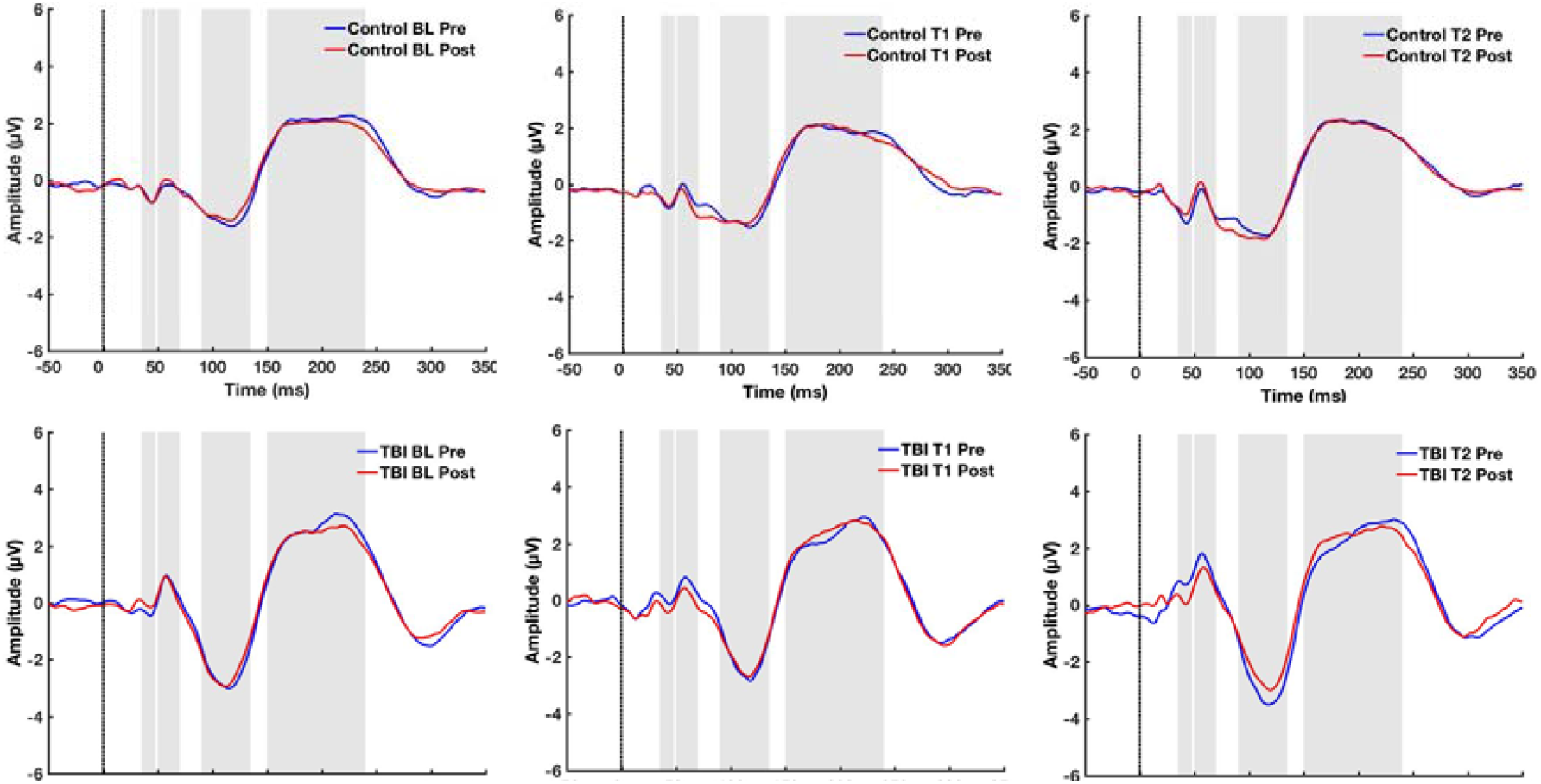
TEP Grand Average waveforms for Control and mTBI participants at each time point. The four peaks of interest and their time windows are highlighted (sequentially from left to right: N45, P60, N100 then P200), with the TMS pulse indicated with a line.

#### Within Group Comparisons -mTBI Group

First, we conducted within-group comparisons on the amplitude of TEP’s pre-and post iTBS at each time point. In the mTBI group, at the sub-acute timepoint, we found that iTBS induced a reduction of the N45 TEP component amplitude (the amplitude became less negative) (*N* = 29, *p* = 0.022). Differences were greatest in the fronto-central region (see Figure 3A). At 3-month follow up, no effect of iTBS on TEP amplitude was demonstrated (*N* = 20, *p* > 0.025). At 6-month follow up, iTBS induced an increase in amplitude of the P200 TEP component in mTBI participants (*N* = 13, *p =* 0.016). Differences were greatest in the left frontal region (see Figure 3B).

**Figure 3.**
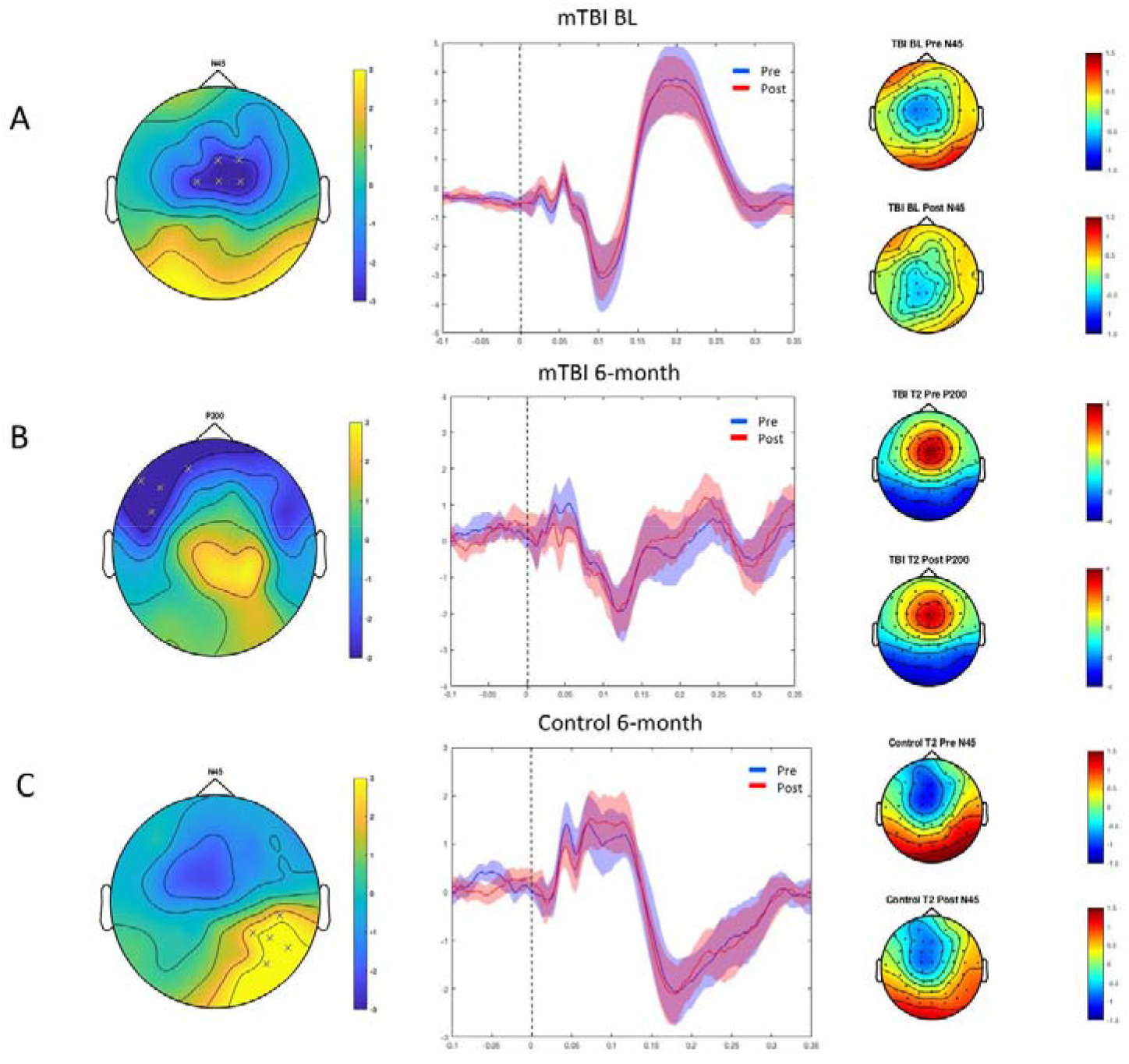
A— Adjacent electrodes (*p* < 0.025) clustering in the central region when testing for an N45 component in the latency range from 35-50 ms post TMS pulse between Pre_iTBS and Post_iTBS conditions in mTBI participants at BL. B— Adjacent electrodes (*p* < 0.025) clustering in the left frontal region when testing for an P200 component in the latency range from 150-240ms post TMS pulse between Pre_iTBS and Post_iTBS conditions in mTBI participants at 6 month follow up. C— Adjacent electrodes (*p* < 0.025) clustering in the right parieto-occipital region when testing for an N45 component in the latency range from 35-50 ms post TMS pulse between Pre_iTBS and Post_iTBS conditions in control participants at 6 month follow up. In A-C, middle graphs display the TEP waveform recorded over the EEG electrodes identified below the critical primary alpha level. Shaded areas indicate 95% confidence intervals. The topoplots on the left show the distribution of activity between conditions (Pre_iTBS to Post_iTBS) for each component.

#### Within Group Comparisons - Control Group

At the sub-acute timeopint (*N=* 26) and at 3-month follow up (*N= 25)* iTBS did not induce amplitude differences for any TEP component in control participants (*p* > 0.025). At 6-month follow up, iTBS induced a change in the N45 TEP component (*N*= 23, *p =* 0.015). Differences were greatest in the right parietal region, which showed positive voltages at both time points, but smaller positive amplitudes following iTBS (see Figure 3C).

#### Between Group Comparisons

Next, we performed between-group comparisons at each time point on the amplitude of TEP’s using difference scores (Δ = Post_iTBS – Pre_iTBS). Testing for an effect in each peak of interest in the pre-defined latency ranges, cluster-based permutation tests revealed no significant differences between control and mTBI participants at the sub-acute timepoint or 3-month follow up at any TEP component (p > 0.025). However, the effect of iTBS was significantly different between mTBI and control participants for the ΔN45 component at 6-month follow up. Inspection of waveforms indicated that in the left frontal region iTBS resulted in a larger amplitude N45 (more negative) in mTBI participants and a smaller amplitude N45 (less negative) in controls. In the right parieto-occipital region, results were in the opposite direction, iTBS resulting in smaller amplitude N45 (less negative) in mTBI participants and a larger amplitude N45 (more negative) in controls. In both brain regions, the iTBS induced difference was larger in mTBI participants (see Figure 4).

**Figure 4.**
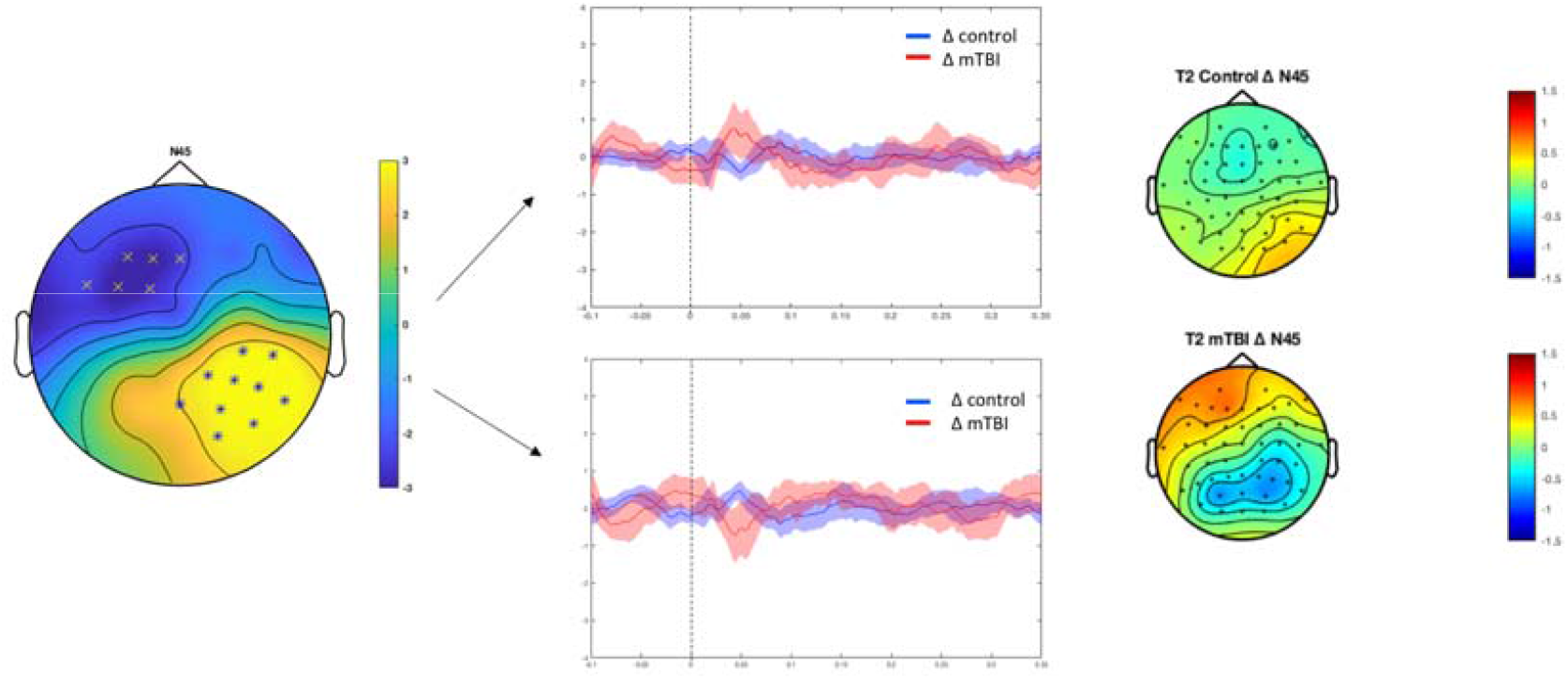
Comparison of the change in transcranial magnetic stimulation (TMS)-evoked N45 amplitude between control and mTBI participants at 6-month follow up. The line graph demonstrates the Post_iTBS minus Pre_iTBS difference TEP waveform recorded over the EEG electrodes identified below the critical primary alpha level. Shaded areas indicate 95% confidence intervals. The topoplots on the right show the distribution of change in Pre_iTBS to Post_iTBS activity for each group for the Δ N45 TEP component.

### Working memory neurophysiology and cognitive performance

There was no effect of iTBS on neural response strength (GFP), topography (TANOVA) or working memory performance (digit span accuracy) at any time point for either mTBI or controls (*p* > 0.05). Mean digit span accuracy is presented in Figure 5.

**Figure 5.**
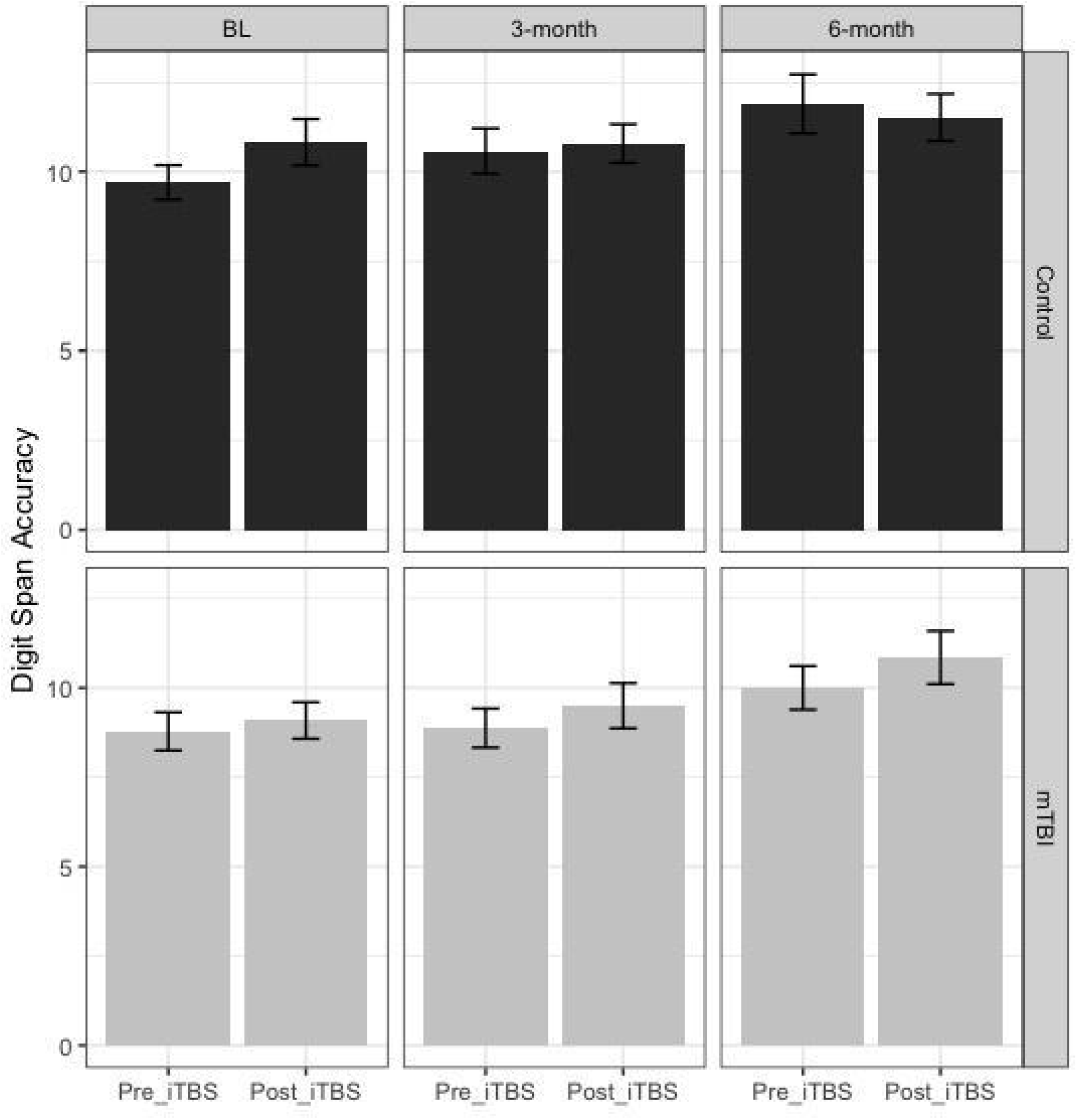
Mean and SD for the control (black) and mTBI (grey) groups for digit span accuracy.

## 4. Discussion

To the best of our knowledge, this is the first study to investigate the effects of iTBS on neurophysiology and cognitive performance post mTBI and across recovery. Overall, results were not consistent with our hypotheses. Firstly, the consistency of iTBS induced changes in neurophysiology (as measured by TEP’s and revealed by comparisons across the time points) was poor, limiting our interpretation of observed changes at specific time points. Secondly, iTBS did not significantly modulate neurophysiology during a working memory task nor behavioral working performance at any time point. Possible explanations for the observed pattern of results are discussed.

Our primary aim was to investigate how mTBI participants responded to the plasticity inducing protocol, iTBS. We first hypothesised that injury induced pathophysiology would dysregulate the brain’s neuroplastic potential, as indexed by a lack of response to iTBS. At the sub-acute timepoint (within one month post injury for mTBI participants), within group analyses showed iTBS resulted in a smaller N45 (less negative) component in fronto-central regions. While TEPs (particularly later TEPs) have been demonstrated to be influenced by somatosensory and auditory processing of the TMS pulse (Conde et al., 2019; Nikouline et al. 1999; Biabani et al. 2019; Biabani et al. 2021), TMS studies in the motor cortex have also associated the N45 with GABAA-mediated inhibition (Premoli et al., 2014) and we have previously reported an increased N100 amplitude in mTBI participants compared to controls, a TEP that has also been associated with inhibitory processes (Coyle et al, 2022a,b). As such, changes to the N45 in response to iTBS reflect changes to cortical reactivity, and may reflect changes to inhibitory signaling in response to excitatory stimulation (i.e. iTBS), perhaps reflecting increased neuroplastic sensitivity in the sub-acute phase post injury. Our second hypothesis was that these changes would normalise across time. No iTBS induced differences were demonstrated at the 3-month follow up, possibly suggesting that the increased neuroplastic sensitivity in the mTBI group (as indexed by N45 amplitude changes at the sub-acute timepoint) may have resolved. However, at 6 months’ post injury iTBS was shown to result in a larger amplitude (more positive) P200 TEP component in the left frontal region of the mTBI group. The physiological origin of the P200 TEP is not well established and the factors that may have contributed to this finding in the late stage of recovery from mTBI unclear.

In control participants, we expected to see changes to neurophysiology in response to iTBS that was consistent across time points. The absence of any significant cluster-based changes following iTBS at the sub-acute timepoint and 3-month follow up suggested that using the current protocol iTBS did not modulate cortical reactivity. However, a larger amplitude (more negative) N45 TEP component in the right parieto-occipital region at the 6-month follow up was also demonstrated. To our knowledge, N45 changes in control participants in response to iTBS have not previously been reported. Notably, we also did not replicate previous work demonstrating modulations in N100 or P200 amplitude in response to iTBS in healthy controls (Chung et al., 2017; Chung, Rogasch, Hoy, Sullivan, et al., 2018; Chung et al., 2018). This finding indicates we were unable to establish consistency of response to iTBS across time in healthy participants.

Lastly, we investigated if control and mTBI participants responded differently to iTBS over the course of recovery. At the sub-acute timepoint and 3-month follow up results demonstrated no between-group differences. However, at 6-month follow up, there was a significant group difference in Δ N45 amplitude in response to iTBS. Inspection of graphs suggested frontally N45 amplitude decreased (became more negative) in controls, whilst N45 amplitude increased (became more negative) in mTBI participants following iTBS. In the posterior region, results were in the opposite direction, N45 amplitude increased (became more negative) in controls, whilst N45 amplitude decreased (became less negative) in mTBI participants. Overall, the difference induced by iTBS was largest in mTBI participants. The factors that may have contributed to the divergent effect of iTBS at a late stage in recovery are unclear, and possible limitations to our experimental design are discussed below.

In contrast to previous research demonstrating improved cognitive performance after iTBS, we found that iTBS did not induce changes in digit span accuracy, neural response strength or the topographical distribution of neural activity in either control or mTBI participants at any time point. There are several possible explanations for this. Firstly, our cognitive task design may have influenced our ability to show differences following iTBS. A step-wise approach to the digit span task was chosen to reduce individual variation in cognitive performance and enable measurement of participants with different working memory capacities. However, our task design may have increased variability in the data. Individuals with lower working memory capacity had a smaller number of available epochs, as two consecutive errors resulted in cessation of the task. iTBS may have induced larger performance gains in lower capacity individuals (Guse, Falkai, & Wobrock, 2010), however this effect could have been averaged out by individuals with higher capacities and a greater number of epochs. Our measure of neural response strength, global field power (GFP), is widely used in neurophysiology as a parameter reflecting the strength of total underlying brain activity. The finding of no differences in either the GFP comparison or the topographical distribution of activity from pre-to post iTBS also suggests iTBS did not modulate activity in the stimulated area or interconnected regions during working memory task. The lack of changes in cognitive performance, GFP or topographical distribution of neural activity highlights the fact that our understanding of the effects of iTBS on cognition and plasticity remains incomplete.

Several limitations should be noted. Our study had a relatively small sample with attrition over time. Different sample sizes at each time point prevented us from completing repeated measures analyses, which increased the multiple comparisons in our study, reducing power and possibly introducing additional variability into our data set. This is a limitation, as a combination of both extrinsic and intrinsic factors contribute to variability in neurophysiological and behavioural response to NIBS protocols (Huang et al., 2017). Firstly, study design and stimulation protocol parameters contribute to mixed results in response to NIBS protocols (Huang et al., 2017; Lopez-Alonso et al., 2014). The current study examined the immediate effects of a single dose of iTBS at 70% of RMT. However, delayed rather than immediate after-effects may better reflect the modulatory effect of iTBS on network activity (Huang et al., 2017) and working memory performance following NIBS has demonstrated greater effect sizes 30-40 minutes post stimulation compared to immediately following stimulation (Chung, Rogasch, Hoy, & Fitzgerald, 2018; Hill, Rogasch, Fitzgerald, & Hoy, 2017). It has also been suggested that iTBS is dose-dependent and repeated application may be of benefit (Nettekoven et al., 2014). Lastly, the effect of stimulation intensity may have contributed. Recent research suggesting iTBS at 75% of RMT results in maximal neurophysiologic change (Chung et al., 2018). iTBS at 90% vs 110% of RMT to the DLPFC has also been shown to result in different functional connectivity activation patterns (Alkhasli, Sakreida, Mottaghy, & Binkofski, 2019). As such, the temporal window post iTBS, single dose, and choice of stimulation intensity may have mediated our findings. Secondly, well controlled studies have demonstrated inherent inter and intra individual variability in the response of neuronal circuits that influence neuroplasticity induction in response to NIBS (Ridding & Ziemann, 2010). Regarding inter and intra individual variability, we assessed mean response for each group. Recent studies have classified participants based on their response to NIBS (e.g. expected responder, non-responder and opposite responder) (Nakamura et al., 2016; Simeoni et al., 2016) to assess the directionality of stimulation effects and minimize the effects of variability on results. Future research that uses a similar approach to classify mTBI participants may be of benefit, especially when assessing cognitive effects. Thirdly, the nature of mTBI pathophysiology may further complicate investigation of plasticity processes. One of the key challenges in mTBI diagnosis and prognosis is heterogeneity, and it has been proposed that the diffuse and dynamic nature of mTBI may make each injury potentially unique (Bigler & Maxwell, 2012; Coyle et al., 2018). Consequently, mTBI induced pathophysiology may further increase inter and intra individual variability in response to NIBS compared to healthy subjects. For example, a recent paper that combined NIBS with neuroimaging demonstrated white matter integrity within a stimulated network influences the behavioral response to stimulation (Li et al., 2019). This suggests structural connectivity may mediate response to stimulation and an individualized approach to NIBS parameters may be necessary in mTBI. Fourthly, although the primary aim of the paper was to investigate the effects of iTBS on neurophysiology, and explore concurrent neurocognitive effects, we acknowledge the absence of sham reduces our ability to conclude any observed changes were iTBS induced, as other variables may have had an influence.

In the current study, iTBS did not reliably modulate neurophysiology or cognitive performance in mTBI participants or controls. No clear relationship between iTBS modulated TEP components and recovery was demonstrated in mTBI and poor consistency across time points in the control group limited interpretation of findings in mTBI. Regarding cognitive performance, our findings did not replicate previous iTBS induced working memory improvements in controls, and there was no evidence to suggest iTBS modulated digit span accuracy in our mTBI sample. Study design factors and inter and intra individual variability in response to NIBS may explain our inconsistent findings. Our results should be interpreted with acknowledgement of the study limitations. The mechanisms and principles that underlie neuroplastic recovery following mTBI remain an area of significant interest, however more work is needed to understand the effects of iTBS on plasticity and cognition prior to therapeutic application in a mTBI sample.

## Data Availability

We do not have ethical approval to make this data publicly available, as our approval predated our inclusion of such approvals (which we now do routinely).

